# Guided antiplatelet therapy with P2Y12 antagonists in patients undergoing percutaneous coronary intervention: 3 systematic reviews with meta-analyses of randomized controlled trials with homogeneous design

**DOI:** 10.1101/2023.05.25.23290520

**Authors:** Simone Birocchi, Matteo Rocchetti, Alessandro Minardi, Gian Marco Podda, Alessandro Squizzato, Marco Cattaneo

## Abstract

**Background:** Conflicting results were reported by randomized controlled trials (RCTs) exploring guided therapy (GT) with anti-P2Y12 drugs in patients undergoing percutaneous coronary intervention (PCI). Meta-analyses of RCTs failed to clearly identify what GT strategy, if any, is effective, because they lumped together RCTs with heterogeneous designs, comparing either genotype-GT or platelet function test (PFT)-GT with unguided standard therapy. Some meta-analysis also included RCTs that did not actually explore GT, but tested the effects of switching patients with high on-treatment platelet reactivity (HTPR) to alternative therapies (HTPR-Therapy), rather than comparing GT with unguided standard therapy. We performed 3 distinct systematic reviews with meta-analyses, each exploring only RCTs with homogeneous design.

**Methods:** MEDLINE, Embase and Central databases were searched for RCTs testing genotype-GT, PFT-GT or HTPR-Therapy in PCI-treated patients, through October 1^st^ 2022. Two reviewers extracted the data. Risk ratios (RR) (95% confidence intervals) were calculated. Primary outcomes were major bleedings (MB) and major adverse cardiovascular events (MACE).

**Results:** In 7 genotype-GT RCTs, RR were: MB, 1.06 (0.73–1.54; p=0.76); MACE, 0.64 (0.45–0.91; p=0.01), but significant risk reduction was observed only in RCTs performed in China (0.30, 0.16-0.54; p<0.0001) and not elsewhere (0.74, 0.46-1.18; p=0.21). In 6 PFT-GT RCTs, RR were: MB, 0.91 (0.64-1.28, p=0.58); MACE, 0.82 (0.5 –1.19; p=0.30): 0.62 (0.42-0.93; p=0.02) in China, 1.08 (0.82-1.41; p=0.53) elsewhere. In 8 HTPR-Therapy RCTs, RR were: MB, 0.71 (0.41-1.23; p=0.22); MACE, 0.57 (0.44–0.75; p<0.0001): 0.56 (0.43-0.74, p<0.0001) in China, 0.58 (0.27-1.23, p=0.16) elsewhere.

**Conclusion:** No GT strategy affected MB. Genotype-GT but not PFT-GT reduced MACE; subgroup analysis revealed that genotype-GT and PFT-GT reduced MACE in China, but not elsewhere. PFT-GT (which analyzed both patients with and without HTPR) performed poorly compared to HTPR-Therapy (which analyzed HTPR patients only), likely due to inaccurate identification of HTPR patients by PFTs. PROSPERO registration: CRD42022362739.

## INTRODUCTION

Patients treated by percutaneous coronary intervention (PCI) are treated with dual antiplatelet therapy (DAPT) with aspirin plus an antagonist of the P2Y12 platelet receptor for adenosine diphosphate (ADP), to reduce the risk of major adverse cardiovascular events (MACE) (1-4). Clopidogrel, the most commonly used anti-P2Y12 drug, does not adequately inhibit platelet function in about 1/3 of treated patients, who display high on-treatment platelet reactivity (HTPR) (5-8). Variability in the generation of the active metabolite of clopidogrel, which is a pro-drug, is the main cause of inter-individual variability of pharmacological response (6,8). Loss of function (LOF) and gain of function (GOF) mutations of cytochrome-P450 (CYP) and the homozygous 3435C→T mutation of ABCB1, a gene encoding for the efflux pump P-glycoprotein (8-12), are associated with response variability to clopidogrel, in combination with other variables, some of which are yet unknown (13). Meta-analyses of observational studies demonstrated that HTPR in clopidogrel-treated patients is associated with inadequate protection from thrombotic events (14-18). Two alternative oral anti-P2Y12 drugs, prasugrel and ticagrelor, display more favorable pharmacokinetics than clopidogrel, which confers more consistent and efficient inhibition of platelet function and thrombosis, resulting in net clinical benefit compared to clopidogrel (19-21).

Randomized controlled trials (RCTs) have been designed to test whether tailoring anti-P2Y12 treatment to individual patients would increase the clinical benefit. Therapy was guided based on either the patients’ genotypes (genotype-GT) or the results of platelet function tests (PFT-GT) performed after the start of standard therapy (Table 1). In most published RCTs, patients on clopidogrel who had been randomized to the testing arm at enrolment were escalated to potentially more effective therapies when they displayed LOF mutations or HTPR. Few RCTs explored the efficacy and safety of guided de-escalation to clopidogrel of patients on prasugrel or ticagrelor displaying normal genotypes or very low on-treatment platelet reactivity (LTPR) (24,33). In these RCTs, patients were randomized to guided therapy (GT) or unguided standard therapy and the safety and efficacy of the two strategies were compared. It must be noted, however, that some published RCTs based on PFT in clopidogrel-treated patients had a different design from the above: all patients underwent testing and only those with HTPR were randomized to continue on clopidogrel or switch to potentially more effective drugs (35-42). This last group of RCTs, therefore, does not actually test the safety and efficacy of GT (which should be compared to those of unguided therapy), but evaluates the safety and efficacy of alternative therapies for patients with HTPR on clopidogrel (HTPR-Therapy) (Table 1).

**Table 1.** Main characteristics of the randomized controlled trials included in the 3 systematic reviews and meta-analyses

| STUDY | NUMBER OF PATIENTS (MEN) | COUNTRIES | ACS / PCI (%) | GENE VARIANT / TYPE OF TEST (HPR threshold) | PATIENTS WITH MUTANT GENOTYPE / HTPR or LTPR † | STANDARD ANTIPLATELET THERAPY ‡ | ALTERNATIVE ANTIPLATELET THERAPY ‡ | DURATION OF THERAPY / FOLLOW-UP (months) | Definition of MACE | Definition of MAJOR BLEEDING |
| --- | --- | --- | --- | --- | --- | --- | --- | --- | --- | --- |
| **1 - RCTs EVALUATING GUIDED ANTI-P2Y12 THERAPY** |  |  |  |  |  |  |  |  |  |  |
| **1a - Genotype-guided anti-P2Y12 therapy** |  |  |  |  |  |  |  |  |  |  |
| **Xie (22)** | 600 (468) | China | 100 / 100 | CYP2C19*2<br>CYP2C19*3 | LOF 52.5% | Clopidogrel<br>75 mg | Clopidogrel<br>150 mg (42.5%);<br>Clopidogrel<br>150mg +<br>Cilostazol<br>100 mg bid (10%) | 6 / 6 | Death from any cause; MI; stroke; ischemia-driven TVR | NA |
| **Notarangelo (23)** | 888 (605) | Italy | 96.8 / 62.2 | ABCB1 3435T<br>CYP2C19*2<br>CYP2C19*17 | 3435 TT 26.4 %<br>LOF 33.5%<br>GOF 39.1% | Clopidogrel<br>75 mg (50.7%);<br>Prasugrel<br>5 mg (8.4%);<br>Ticagrelor<br>90 mg bid (32.7%) | Clopidogrel<br>75 mg (43.3%);<br>Prasugrel<br>5 mg (7.6%);<br>Ticagrelor<br>90 mg bid (42.6%) | 12 / 12 | CV death; first occurrence of non-fatal MI; non-fatal stroke; ST | NA |
| **Claassens (24)** | 2488 (1862) | Netherland<br>Belgium<br>Italy | 100 / 100 | CYP2C19*2<br>CYP2C19*3 | LOF 31.3% | Ticagrelor<br>90 mg bid, or<br>Prasugrel<br>5 mg (92.1%);<br>Clopidogrel<br>75 mg (7%) | Ticagrelor<br>90 mg bid or<br>Prasugrel<br>5 mg (39.1%);<br>Clopidogrel<br>75 mg (60.6%) | 12 / 12 | CV death; ACS; definite ST; stroke; urgent TVR | BARC 3 or 5 |
| **Pereira (25)** | 5302 (3983) | USA<br>Canada<br>Mexico<br>South Korea | 82 / 100 | CYP2C19*2<br>CYP2C19*3 | LOF 34.2% | Clopidogrel<br>75 mg | Ticagrelor<br>90 mg bid | 12 / 12 | CV death, MI, stroke, definite or probable ST, severe recurrent ischemia | TIMI bleeding criteria |
| **Tuteja (26)** | 504 (307) | USA | 51 / 100 | CYP2C19*2<br>CYP2C19*17 | LOF 27.0%<br>GOF 36.0% | Clopidogrel<br>75 mg (78.8%);<br>Prasugrel<br>5 mg (11%);<br>Ticagrelor<br>90 mg bid (10%) | Clopidogrel<br>75 mg (70.1%);<br>Prasugrel<br>5 mg (16.1%);<br>Ticagrelor<br>90 mg bid (13.3%) | NA / 16 | First MI, ischemic CV death, stroke, ST, urgent TVR | BARC 3 or 5 |
| **Al-Rubaish (27)** | 687 (555) | Saudi Arabia | 100 / 100 | CYP2C19*2 | LOF 31.5% | Clopidogrel<br>75 mg | Ticagrelor<br>90 mg bid | 12 / 12 | Death; MI; ST | PLATO bleeding criteria |
| **Shi (28)** | 301<br>(226) | China | 100 /<br>100 | CYP2C19*2<br>CYP2C19*3 | LOF 57.3% | Clopidogrel<br>75 mg (75.5%);<br>Ticagrelor 90 mg<br>bid (25%) | Clopidogrel<br>75 mg (42,8%);<br>Ticagrelor 90 mg<br>bid (57,2%) | NA / 12 | All-cause death; MI;<br>stroke; urgent TVR;<br>ST | NA |
| **1b - PFT-guided anti-P2Y12 therapy** |  |  |  |  |  |  |  |  |  |  |
| **Collet (29)** | 2440<br>(1970) | France | 26.6 /<br>100 | VerifyNow<br>(>235 PRU) | HPR 15,6% | Clopidogrel<br>75 mg (93.5%);<br>Prasugrel<br>5 mg (5.8%) | Clopidogrel<br>75 mg (47%);<br>≥150 mg (43%);<br>Prasugrel<br>5/10 mg (9.3%) | 12 / 12 | Death from any<br>cause; MI; ST;<br>stroke; TIA; TVR | STEEPLE<br>bleeding criteria |
| **Hazarbasanov (30)** | 192<br>(126) | Bulgaria | 54 /<br>100 | Multiplate<br>analyzer (>46 U) | HPR 18.5% | Clopidogrel<br>75 mg | Clopidogrel<br>150 mg | 6 / 6 | Cardiac death; MI;<br>ST; ischemic stroke | TIMI<br>bleeding criteria |
| **Li X (31)** | 384<br>(289) | China | 100 /<br>100 | TEG<br>(IPA<30%) | HPR 40,4% | Clopidogrel<br>75 mg | Clopidogrel<br>150 mg | 6 / 6 | MI; ST; death;<br>emergency TVR | NA |
| **Zhu (32)** | 305<br>(201) | China | 100 /<br>100 | LTA<br>(IPA<10%) | HPR 26,6% | Clopidogrel<br>75 mg | Clopidogrel<br>75 mg +<br>Cilostazol<br>100 mg bid | 12 / 12 | CV death; MI; TVR;<br>stroke | TIMI<br>bleeding criteria |
| **Cayla (33)** | 877<br>(533) | France | 100 /<br>100 | VerifyNow<br>(>207 PRU) | HPR 4%<br>LPR 42% | Prasugrel<br>5 mg (93%)<br>10 mg (1%);<br>Clopidogrel<br>75 mg (4%) | Prasugrel<br>5 mg (55%)<br>10 mg (4%);<br>Clopidogrel<br>75 mg (39%) | 12 / 12 | CV death; MI; ST;<br>urgent TVR | TIMI<br>bleeding criteria |
| **Zheng (34)** | 2285<br>(1904) | China | 0 /<br>100 | PL-12 §<br>(>55% MAR) | HPR 62,8% | Clopidogrel<br>75 mg | Ticagrelor<br>90 mg bid | 6 / 6 | CV death; MI; ST;<br>urgent TVR; stroke | BARC 3 or 5 |
| **2 - RCTs EVALUATING THERAPY OF HTPR ON CLOPIDOGREL** |  |  |  |  |  |  |  |  |  |  |
| **Price (35)** | 2214<br>(1441) | USA<br>Canada | 39.8 /<br>100 | VerifyNow<br>(>230 PRU) | 100% | Clopidogrel<br>75 mg | Clopidogrel<br>150 mg | 6 / 6 | CV death;<br>non-fatal MI; ST | GUSTO<br>bleeding criteria |
| **Ari (36)** | 94<br>(72) | Turkey | 0 /<br>100 | VerifyNow<br>(<40% IPRU) | 100% | Clopidogrel<br>75 mg | Clopidogrel<br>150 mg | 1 / 6 | CV death; MI; ACS;<br>TVR; ST | TIMI<br>bleeding criteria |
| **Trenk (37)** | 423<br>(307) | Germany<br>USA | 0 /<br>100 | VerifyNow<br>(>208 PRU) | 100% | Clopidogrel<br>75 mg | Prasugrel<br>10 mg | 6 / 6 | CV death; MI;<br>stroke; cardiac re-<br>admission | TIMI<br>bleeding criteria |
| **Samardzic (38)** | 87<br>(49) | Croatia | 100 /<br>100 | Multiplate<br>analyzer (>46 U) | 100% | Clopidogrel<br>75 mg | Clopidogrel<br>≥150 mg | 12 / 12 | Death; non-fatal<br>MI; TVR;<br>ischemic stroke | BARC 3 or 5 |
| **Li Y (39)** | 840<br>(555) | China | 100 /<br>100 | LTA<br>(PA>55%) | 100% | Clopidogrel<br>75 mg | Clopidogrel<br>75/150 mg +<br>Cilostazol<br>50/100 mg bid | 12 / 12 | All-cause death;<br>non-fatal MI; TVR;<br>stroke | TIMI<br>bleeding criteria |
| **Tang (40)** | 721<br>(433) | China | 59.7 /<br>100 | TEG<br>(MA<sub>ADP</sub>>47mm +<br>ADP-IPIR<50%) | 100% | Clopidogrel<br>75 mg | Clopidogrel<br>150 mg | 12 / 18 | All-cause death; MI;<br>TVR; stroke | NA |
| **You (41)** | 300<br>(195) | China | 100 /<br>100 | Platelet VASP-P<br>(≥50% PRI) | 100% | Clopidogrel<br>75 mg | Ticagrelor<br>90 mg bid | 12 / 12 | All-cause death; CV<br>death; recurrent<br>MI; TVR;<br>ischemic stroke | NA |
| **Ying (42)** | 434<br>(156) | China | 84.5 /<br>100 | LTA<br>(PA>40%) | 100% | Clopidogrel<br>75 mg | Clopidogrel<br>150 mg (33.6%);<br>Clopidogrel 75 mg<br>+ Cilostazol<br>100 mg bid<br>(32.7%);<br>Ticagrelor<br>90 mg bid (33.6%) | 12 / 12 | CV death;<br>non-fatal MI;<br>ischemic stroke;<br>TVR; ST;<br>cardiac readmission | TIMI<br>bleeding criteria |

† Guided Therapy Group only. Percentages of prevalence of mutant CYP genotype include both heterozygous and homozygous variants CYP2C19\*2/CYP2C19\*3 (LOF) and CYP2C19\*17 (GOF).
‡ For studies (23-25, 29, 33) in which standard and/or alternative treatments were chosen by treating physicians (based on clinical, PFT and genotype criteria) the relative percentages of patients assigned to each treatment in each arm are indicated, when available on the original report. Drugs were administered were once daily, unless otherwise specified.
§ PL-12 measures platelet aggregation in whole blood, based on changes in platelet count of the sample after the addition of the platelet agonist (ADP in this study).
**Abbreviations:** ABCB1=ATP Binding Cassette Subfamily B Member 1; ACS=acute coronary syndromes; ADP-IPIR=ADP-induced platelet inhibition rate; bid=bis in die; CV=cardiovascular; CYP=cytochrome 450; GOF=gain of function; HTPR=high on-treatment platelet reactivity; HR=hazard ratio; IPA=inhibition of platelet aggregation; IPRU=inhibition of P2Y<sub>12</sub> reactions units; LOF=loss of function; LTPR=low on-treatment platelet activity; LTA=light transmission aggregometry; MA<sub>ADP</sub>=maximal amplitude induced by ADP; MACE=major adverse cardiovascular events; MAR=maximum aggregation rate; MI=myocardial infarction; NA=not available; PA=platelet aggregation; PFT=platelet function test; PRI=P2Y<sub>12</sub> reactivity index; PRU= P2Y<sub>12</sub> reactions units; RCT=randomized controlled trial; RR=relative risk; ST=stent thrombosis; TEG= thromboelastography; TVR=target-vessel revascularization; U=units; VASP-P=vasodilator-stimulated phosphoprotein phosphorylation.

Meta-analyses of published studies gave contrasting results, likely due to the heterogeneity of the included studies. Some meta-analyses lumped together RCTs, non-randomized intervention studies and observational studies (43-47). Meta-analyses that considered RCTs only, or analyzed the results of RCTs separately from those of observational studies, lumped together RCTs with heterogeneous designs, testing genotype-GT or PFT-GT (46), PFT-GT or HTPR-Therapy (48), genotype-GT, PFT-GT or HTPR-Therapy (49). Therefore, these meta-analyses are unable to provide the necessary information of what GT strategy, if any, improves the clinical benefit of anti-P2Y12 therapy. Two meta-analyses included genotype-GT RCTs only (50, 51), while none analyzed PFT-GT RCTs only.

It is potentially misleading to combine in a single meta-analysis RCTs testing GT and testing alternative therapies for HTPR, or RCTs exploring genotype-based tests and exploring PFT-based tests, which have different accuracy and precision in the identification of the patients’ features that influence the selection of treatment strategies. Therefore, we elected to perform 3 separate systematic reviews and meta-analyses, each one for each of the 3 different study designs of RCTs. The reason why we analyzed also RCTs on HTPR-Therapy, in addition to genotype-GT and PFT-GT, is that, although they did not actually explore GT, their results could help interpreting the results of PFT-GT RCTs, considering that both types of RCTs are based on PFTs, but in a different experimental setting.

## METHODS

The development of the study protocol, the conduct of the systematic reviews and meta-analyses, the reporting of results are in accordance with the Preferred Reporting Items for Systematic Reviews and Meta-analyses (PRISMA) guidelines. The study protocol was registered on PROSPERO (CRD42022362739).

### Data Sources and Searches

We searched on MEDLINE, Cochrane Central Register of Controlled Trials (CENTRAL) and Embase using the following key concepts: anti-P2Y12 therapy, platelet function test and genetic test. For each key concept, appropriate free-text words and Medical Subject Headings (MeSH) were developed.

### Study selection

RCTs on adult patients requiring DAPT after PCI for acute coronary syndrome (ACS) or chronic coronary disease (CCD), comparing guided versus standard anti-P2Y12 therapy or alternative versus standard therapy for patients with HTPR on clopidogrel and published until October 1^st^, 2022 were eligible. The records identified with our search were screened by two independent reviewers (MR and AM) at first by title and abstract and then by reading the full texts. A hand search was performed on the reference lists of selected articles to include studies that had not been identified by the search strategy. Duplicate publications and studies non reporting relevant data for the pre-specified outcomes were excluded. In case of disagreements, a third reviewer (MC) was responsible for the final decision. Exclusion criteria were the concomitant use of platelet function testing and genetic testing for guiding anti-P2Y12 therapy, duration of treatment and follow-up ≤30 days.

### Data collection/extraction process and analysis

Data were extracted by two independent reviewers using a standardized data abstraction form, developed according to the sequence of variables required from the primary studies. Disagreements in data abstraction between the two reviewers were solved by a third independent reviewer (MC). Risk ratios (RR) and 95% confidence intervals (CIs) were calculated for each outcome. Heterogeneity among studies was quantified using I² statistic and statistical significance was assessed using χ² test (I²>50% or p<0.10 indicating the presence of significant heterogeneity among studies). Meta-analyses of RR were conducted using the DerSimonian and Laird random effects models, irrespective of the presence of clinical and methodological heterogeneity. For all the analyses, a value of p<0.05, two sided, was considered statistically significant. All statistical analyses were performed using Review Manager 5.3.

### Outcomes

The main safety outcome was major bleeding, defined as BARC 3 and 5. The main efficacy outcome was MACE, as defined by each RCT. The secondary efficacy outcomes were myocardial infarction, cardiovascular death, stent thrombosis and ischemic stroke, while the secondary safety outcome was all bleedings.

We performed the following predefined sub-analyses when at least 2 RCTs provided the necessary data: type of alternative therapy in the guided arm, escalation RCTs, de-escalation RCTs, tertiles of prevalence of patients with ACS (0-33%; 34-66%; 67-100%), sample size <200 or >200 per arm, type of PFT used, countries in which RCTs had been performed.

### Quality assessment

Two reviewers assessed the quality of data in included studies with the Cochrane tool for quality assessment. Funnel plots were generated to investigate the risk of publication bias.

### Registration

The study was registered with PROSPERO (CRD42022362739).

## RESULTS

Our systematic literature search identified 3,835 articles, which were reduced to 2695 after duplicates removal; 16 additional records were then retrieved by manual search of the references of relevant articles, for a total of 2,711 records. After screening by title, abstract and full text, we identified 21 RCTs that fulfilled the criteria for inclusion in our analysis (Figure 1): 7 on genotype-GT (22-28), 6 on PFT-GT (29-34), 8 on HTPR-Therapy (35-42). Descriptions of their characteristics are summarized in Table 1. In all included RCTs, the percentage of patients who underwent PCI was 100%, with the only exception of one RCT (23) in which 62.2% underwent PCI: thus, the frequency of PCI was 98.5% in the overall patient populations enrolled in the included RCTs. The majority of PFT-GT and genotype-GT RCTs aimed at escalating patients in the guided arm with (or at risk of) HTPR on clopidogrel to more efficient antiplatelet treatments. Two RCTs (24, 33) aimed at de-escalating to clopidogrel ACS patients in the guided arm with (or at risk of) LTPR on ticagrelor or prasugrel. Two additional genotype-guided RCT escalated or de-escalated anti-P2Y12 treatment based on the presence of LOF or GOF CYP genotypes in patients who had been assigned to clopidogrel, prasugrel or ticagrelor before randomization, based on physicians’ choice (23, 26). We excluded the TROPICAL-ACS RCT (53), which is considered a PFT-guided de-escalation study and was included in some previous meta-analyses (46, 48). The reason for our choice is that TROPICAL-ACS did not compare a guided de-escalation arm with a non-guided de-escalation arm, but re-escalated to the initial treatment with prasugrel patients with HTPR after de-escalation to clopidogrel. However, with the aim of rendering our meta-analysis comparable to others from this point of view, we included the TROPICAL-ACS study in a sub-analysis.

**Figure 1.**
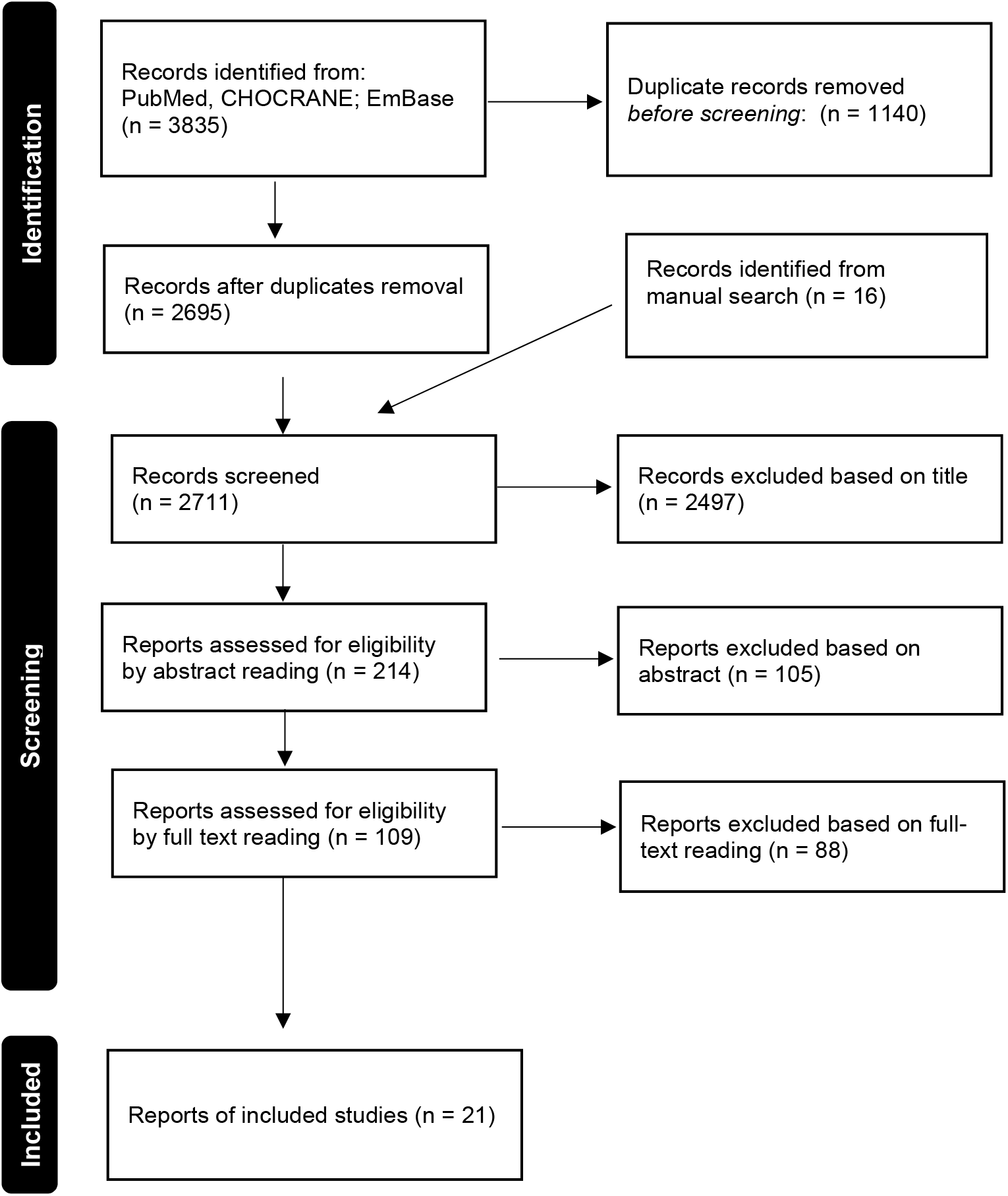
PRISMA flow diagram of the systematic research.

Most included studies were of high quality, with only 4 presenting at least 2 items with high risk of bias (23, 26-28) (Supplementary Figure 1). Funnel plot analysis revealed that the risk of publication bias was low for PFT-GT studies and genotype-GT studies, but potentially significant for HTPR-Therapy studies (Figure 2)

**Figure 2.**
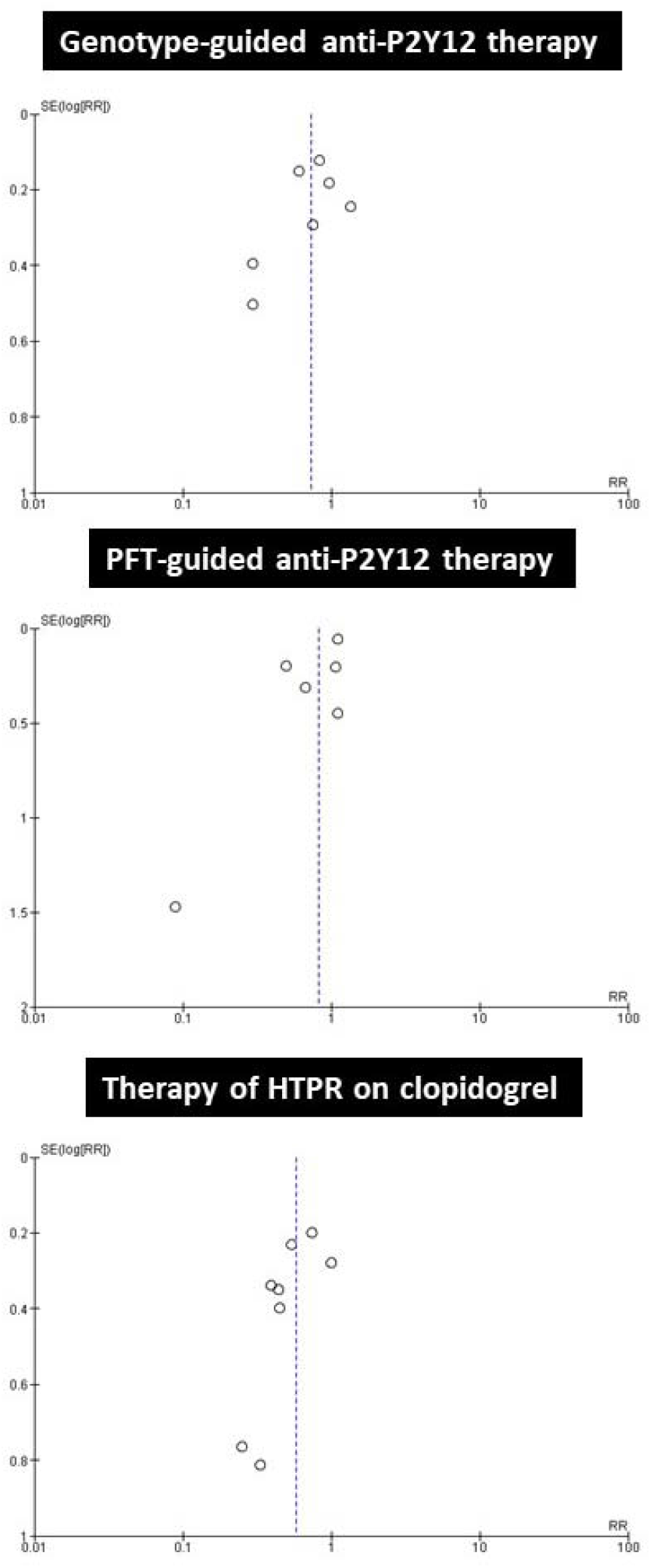
Funnel plots based on the primary efficacy outcome MACE.

## GENOTYPE-GT

### Primary Outcomes

Data on major bleeding were available for 4 studies and a total of 8,955 patients. There was no statistically significant difference in the risk of major bleeding between the genotype-guided group (71 events in 4507 patients, 1.58%) and the standard treatment group (68 events in 4448 patients, 1.53%), RR=1.06 (0.73–1.54; p=0.76) (Figure 3).

**Figure 3.**
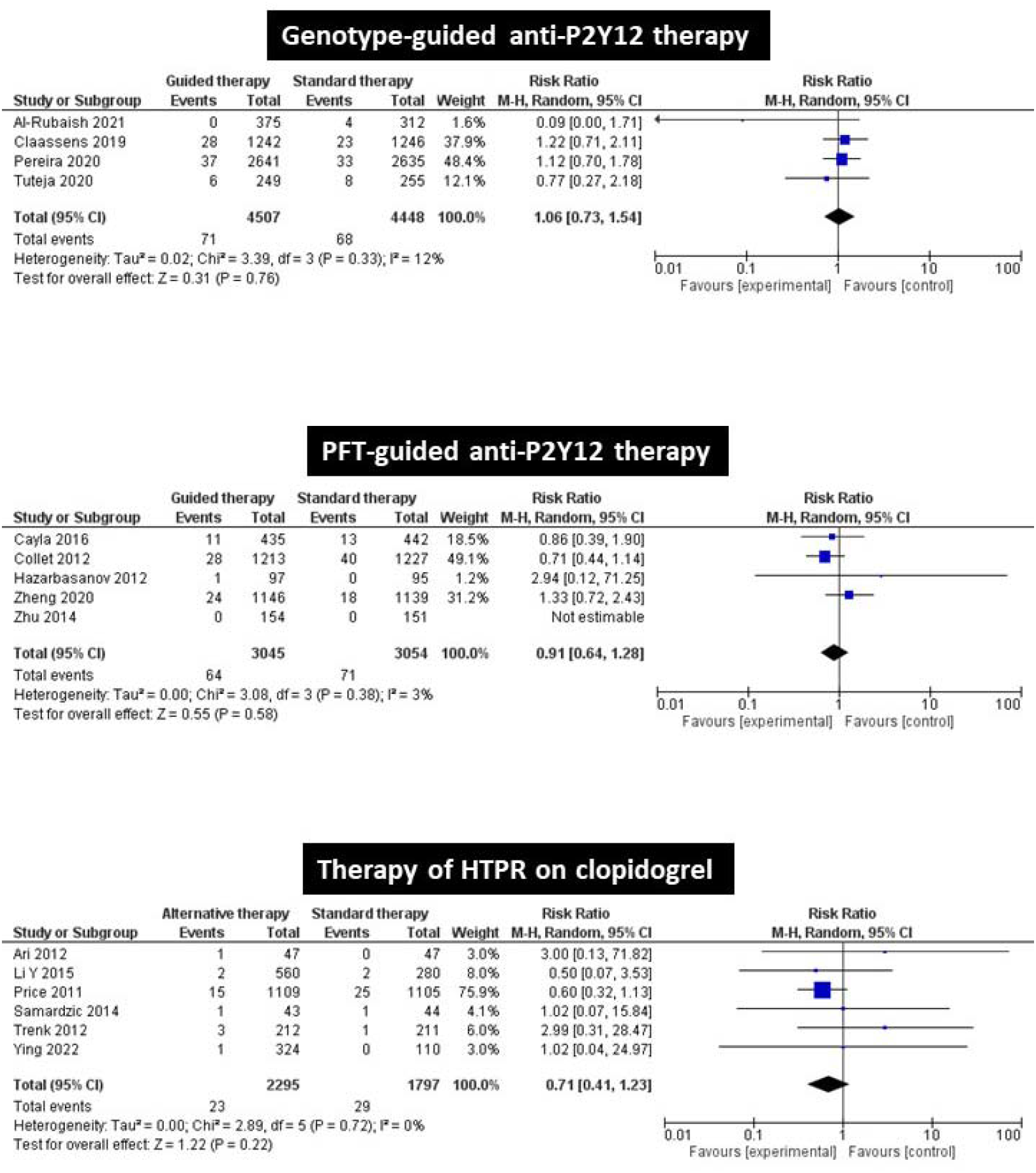
Pooled analysis for the primary safety outcome of Major Bleeding (BARC 3 or 5). Genotype-guided and platelet function tests-guided versus standard anti-P2Y12 therapy; alternative antiplatelet therapies for patients with high platelet reactivity on clopidogrel. PFT= platetel function tests; HTPR= high on-treatment platelet reactivity.

Data on MACE were available for 7 studies and a total of 10,744 patients. The risk of MACE was significantly lower in the genotype-guided group (297 events in 5,457 patients, 5.44%) than in the standard treatment group (398 events in 5,287 patients, 7.52%), RR=0.64 (95% CI, 0.45–0.91; p=0.01) (Figure 4).

**Figure 4.**
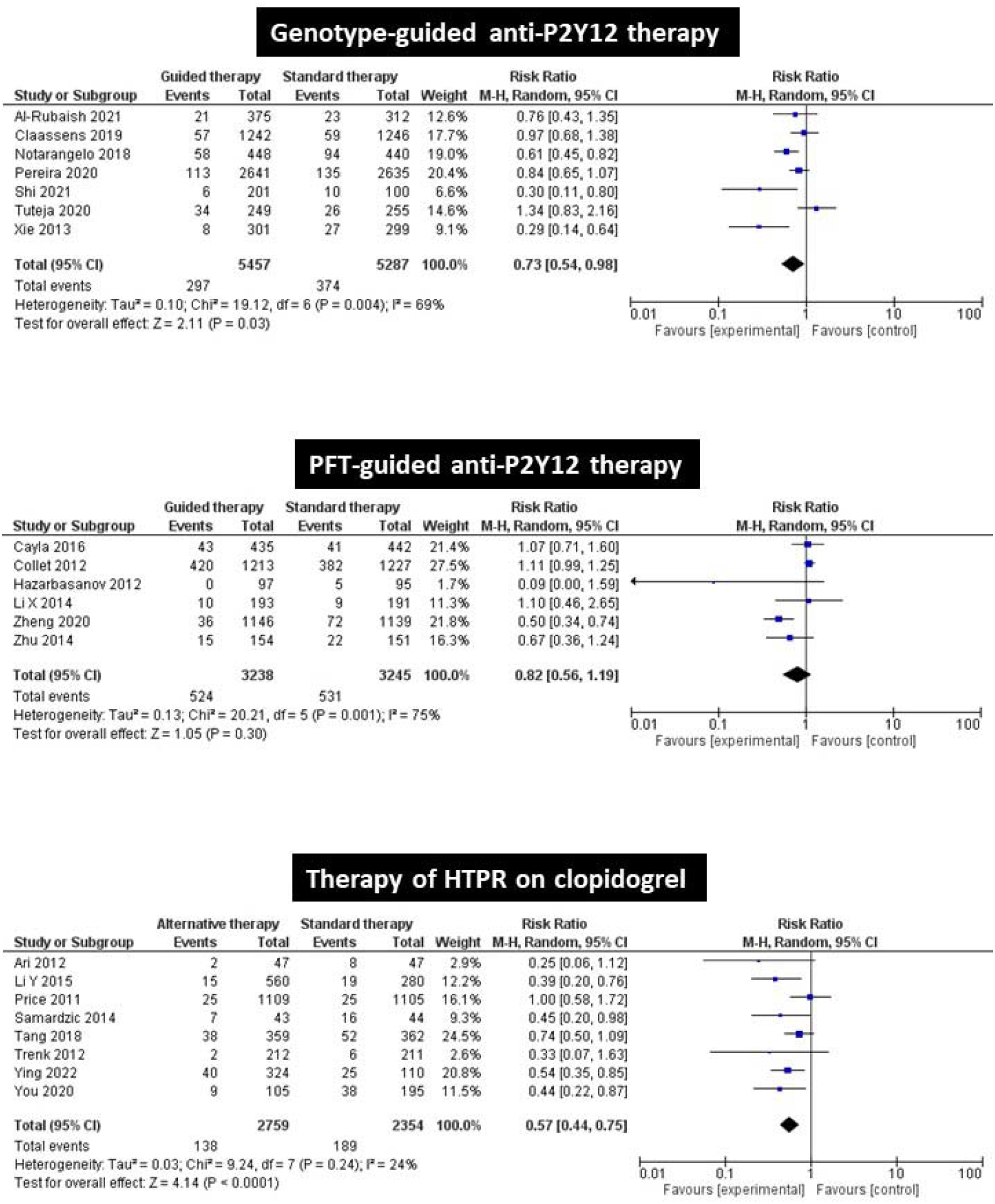
Pooled analysis for the primary efficacy outcome of MACE. Genotype-guided and platelet function tests-guided versus standard anti-P2Y12 therapy; alternative antiplatelet therapies for patients with high platelet reactivity on clopidogrel. PFT= platetel function tests; HTPR= high on-treatment platelet reactivity.

#### Subgroups analyses

The risk of MACE was lower in the genotype-GT group in escalation studies (there were no enough de-escalation studies to be analyzed), in studies that used prasugrel or ticagrelor in genotype-guided arms, in studies with >66% ACS patients (there were not enough studies with lower ACS percentages), and in studies performed in China, while it was similar to non-guided group in RCTs performed in other countries (Table 2), even after exclusion of the study with de-escalation strategy (24) (Supplementary Figures 2,3). All genotype-GT RCTs enrolled >200 patients in each arm.

**Table 2.** Point estimates of Risk Ratios for MACE in the 3 types of randomized controlled trials as a function of the geographical regions in which they had been performed

| Strategy explored in RCTs | RCTs performed in China* |  | RCTs performed outside China* |  |
| --- | --- | --- | --- | --- |
|  | RR, 95% CI<br>(n. of patients) | P value | RR, 95% CI<br>(n. of patients) | P value |
| **Genotype-guided anti-P2Y12 therapy** | **0.30**, 0.16-0.54<br>(901) | <0.0001 | **0.74**, 0.46-1.18<br>(9,869) | 0.21 |
| **PFT-guided anti-P2Y12 therapy** | **0.62**, 0.42-0.93<br>(2,974) | 0.02 | **1.08**, 0.82-1.41<br>(3,509) | 0.53 |
| **Alternative therapies for HTPR on clopidogrel** | **0.56**, 0.43-0.74<br>(2,295) | <0.00001 | **0.58**, 0.27-1.23<br>(2,818) | 0.16 |

\*see Table 1 for details
Abbreviations: MACE=major adverse cardiovascular events; RR= risk ratio; CI= confidence intervals; RCTs= randomized controlled trials; HTPR= high on-treatment platelet reactivity.

### Secondary Outcomes

The risk of myocardial infarction (RR=0.55, 0.42-0.71, p<0.0001) and stent thrombosis (RR=0.67, 0.46-0.96, p=0.03) was significantly lower in genotype-GT, compared to standard therapy. There was no statistically significant difference in the risk of cardiovascular death, ischemic stroke and all bleedings (Supplementary Figure 4).

## PFT-GT

### Primary Outcomes

Data on major bleeding were available for 5 studies and a total of 6,099 patients. There was no statistically significant difference in the risk of major bleeding between the PFT-guided group (64 events in 3,045 patients, 2.1%) and in the standard treatment group (71 events in 3,054 patients, 2.32%), RR=0.91 (0.64-1.28; p=0.58) (Figure 3).

Data on MACE were available for 6 studies, and a total of 6,483 patients. There was no statistically significant difference in the risk of MACE between the PFT-guided group (524 events in 3,238 patients, 16.18%) and the standard treatment group (531 events in 3,245 patients, 16.36%), RR=0.82 (0.5 –1.19; p=0.30) (Figure 4).

#### Subgroups analyses

There were no statistically significant difference in the risk of both MACE and major bleeding between the 2 treatment arms independently of the type of alternative therapy used (higher doses of clopidogrel or other alternative therapies), in escalation strategy studies (there were no enough de-escalation studies to be analyzed), in studies that used VerifyNow as PFT (there were no enough studies using other PFT to be analyzed) and independently of the prevalence of ACS patients (<33% or >66%: there were no enough studies with ACS prevalence 33-66% to be analyzed) (Supplementary Figures 5,6). In contrast, the risk of MACE was significantly lower in the PFT-guided group compared to standard treatment in RCTs performed in China, RR=0.62 (0.42-0.93, p=0.02), while it was similar to that of standard therapy group in RCTs performed in other countries, RR=1.08 (0.82-1.41; p=0.53) (Table 2). There was no statistically significant difference between the 2 treatment arms in the risk of major bleeding both in studies performed in China (RR=1.33, 0.72-2.43; p=0.91) and in other countries (RR=0.76, 0.51-1.14; p=0.19) (Supplementary Figures 5,6). Inclusion of the TROPICAL RCT (53) in our analysis did not substantially change the primary outcome analyses for MACE (RR=0.81, 0.59-1.12, p=0.21) and MB (RR=0.69, 0.66-1.20, p=0.45).

### Secondary outcomes

There was no statistically significant difference in the risk of myocardial infarction, cardiovascular death, stent thrombosis, ischemic stroke and all bleedings between the two treatment arms (Supplementary Figure 7).

## HTPR-THERAPY

### Primary outcomes

Data on major bleeding were available for 6 studies, for a total of 4092 patients. There was a non-significant difference in the risk of major bleeding among patients who were given alternative treatments (23 events in 2295 patients, 1.00%) and among patients who continued on clopidogrel (29 events in 1797 patients, 2.62%), RR=0.71 (0.41-1.23; p=0.22) (Figure 3).

Data on MACE were available for 8 studies, for a total of 5113 patients. The risk of MACE was significantly lower among patients who were given alternative treatments (138 events in 2719 patients, 5.08%) compared to patients who continued on clopidogrel (189 events in 2354 patients, 8.03%), RR=0.57 (0.44–0.75; p<0.0001) (Figure 4).

#### Subgroups analysis

Alternative treatments significantly decreased the risk of MACE in RCTs that used LTA as PFT (RR=0.49, 0.34-0.71; p=0.0002), but not in those that used VerifyNow (RR=0.55, 0.21-1.43; p=0.22). Compared with standard dose clopidogrel-treated patients, the risk of MACE in patients treated with clopidogrel+cilostazol, prasugrel or ticagrelor was significantly lower (RR=0.47, 0.34-0.65; p<0.00001), while it was non-significantly different in patients treated with higher doses of clopidogrel (RR=0.68, 0.45-1.03; p=0.07). The alternative treatments used decreased the risk of MACE independently of the sample size of the studies (<200 o >200) or the prevalence of ACS <33% or >66%, while they were not effective in RCTs including 33%-66% HPR patients with ACS. MACE were similarly lower in the experimental groups in RCTs performed in China (RR=0.56, 0.43-0.74, p<0.00001) and in other countries (RR=0.58, 0.27-1.23, p=0.16), but statistical significance was observed only for RCTs performed in China (Table 2, Supplementary Figures 8,9). The risk of major bleedings was similar in the two treatment arms in all subgroup analyses (Supplementary Figures 8,9).

### Secondary outcomes

The risk of cardiovascular death was significantly lower (RR=0.36, 0.19-0.66, p=0.001) and that of all bleedings was significantly higher (RR=1.23, 1.01-1.50, p=0.04) in the alternative therapies arm, compared to the standard clopidogrel arm. No statistically significant differences were observed in the incidence of myocardial infarction, stent thrombosis and ischemic stroke between the two arms (Supplementary Figure 10).

## DISCUSSION

The value of guiding DAPT with laboratory tests that measure platelet function or identify mutant genotypes that influence the pharmacological response to anti-P2Y12 drugs has long been studied and debated (8,21,54-61). RCTs comparing unguided standard therapy with genotype-GT or PFT-GT have been published together with RCTs that did not actually compare the efficacy and safety of guided therapy, but tested the effects of alternative therapies of patients displaying HTPR on clopidogrel. The results of published RCTs are controversial, partly as a consequence of the small sample size of many of them. Systematic reviews with meta-analyses of RCTs (combined with non-randomized intervention studies and observational studies in some cases) have been performed, which, however, usually lumped together studies with different designs and addressing different questions. Therefore, we elected to perform 3 distinct systematic reviews with meta-analyses, each one being strictly focused on RCTs with homogeneous study question and design.

### Genotype-GT

Genotype-GT did not affect the risk ratio for major bleeding. Although this analysis was restricted to the results of studies performed in western countries, which evaluated this end point, they are compatible with the findings of the 2 RCTs that had been performed in China, in which genotype guidance did not affect the incidence of “any significant bleeding events”.

In contrast, genotype-GT reduced the incidence of MACE. The low RR for MACE (0.57, 0.44-0.75) indicates that the effect of genotype testing is not only statistically significant, but also clinically significant. It must be noted, however, that most of the evidence of protective effects of genotyping stemmed from the two RCTs that had been performed in China (RR=0.30, 0.16-0.54). RCTs performed in other parts of the world failed to show a statistically significant benefit of genotyping (RR=0.74, 0.46-1.18), although they enrolled 10x more patients than RCTs performed in China.

The observation that genotype-GT was efficacious in patients enrolled in China, but not in other countries, is compatible with the marginal role of CYP2C19 mutations in the response to clopidogrel among non-asiatic populations (8,13,57,62), while they could bear greater influence among patients from East/South Asia, in whom the prevalence of CYP2C19 LOF mutations is much higher (63-66). In our systematic review, the prevalence of intermediate (carrying one LOF allele) plus poor metabolizers (carrying two LOF alleles) was 52.5% (22) and 57.3% (28) in RCTs performed in China and 27.0-34.2% in RCTs performed elsewhere, comparably to estimates that were previously reported in large epidemiological studies worldwide (63-66). In addition, it is noteworthy that two meta-analyses showed that the RR for MACE among clopidogrel-treated ACS patients carrying LOF CYP2C19 mutations was higher among patients from East/South Asia, compared to patients from western countries: RR=1.91, 1.61-2.26 *vs* RR=1.23, 1.07-1.40 (67), and RR=2.02, 1.67-2.44 *vs* 1.35, 1.20-1.52 (68). These observations, which suggest that additional factors influence the risk for MACE in chinese carriers of LOF CYPC219 mutations treated with anti-P2Y12 drugs, contribute to account for the greater benefit of genotype-GT among Chinese patients, compared to other patients.

The choice of alternative treatments to clopidogrel 75 mg od for patients with CYP2C19 LOF mutations varied among different RCTs, including ticagrelor, prasugrel, high-doses clopidogrel alone or in combination with cilostazol. Based on the limited data available, we could not identify the most effective alternative treatment in these patients.

### PFT-GT

Our analysis of RCTs of PFT-GT revealed that it had no statistically significant effect on the incidence of both major bleedings and MACE. However, a sub-analysis showed that PFT-GT significantly reduced the incidence of MACE in Chinese patients but not in non-asiatic patients. The explanation for this difference is the same already given for the discrepancy observed in these populations for genotype-GT. Similarly to the prevalence of LOF mutations of the CYP2C19 gene, which was higher among Chinese patients, the prevalence of HPR was higher among clopidogrel-treated Chinese patients (26.6-62.8%) (31,32,34) compared to non-asiatic patients (15.6% and 18.5%) (29,30) (Table 1). It is noteworthy that the statistically non-significant effect of PFT-GT in our global analysis, as opposed to the statistically significant effects of genotype-GT, was observed despite the fact that the relative proportion of enrolled Chinese patients in RCTs of PFT-GT was much higher (45.8%) than in genotype-GT RCTs (8.4%). Although this is an indirect evidence, these data suggest that PFT-GT is less effective than genotype-GT both among Chinese patients (RR=0.63 vs 0.30) and among patients from other countries (RR=1.08 vs 0.74).

The lower efficacy of PFT-GT compared with genotype-GT is to be found in the unsatisfactory diagnostic accuracy of PFT and the lability of the platelet reactivity phenotype, which may switch from HPR to normal platelet reactivity or LPR and back to HTPR over time in a high proportion of patients (69-74), which contrasts with the stability of the CYP2C19 genotype. The lability of the platelet reactivity phenotype reflects the physiological variability of platelet function with time, but is also due to the lack of standardization of pre-analytical variables, including the time of the day of blood sampling [platelet function follows a circadian rhythm (75,76)], the time elapsed since the acute event and since the last drug intake (8). Moreover, the agreement among different PFT evaluating the response to anti-P2Y12 therapy is very low (77-81), thus implying that the diagnostic accuracy of the tests is poor. The inaccuracy of PFT in identifying HTPR patients accounts for their failure to improve the clinical outcomes of clopidogrel-treated patients in RCTs, despite their ability to document the association of HTPR with poor clinical outcomes in observational studies of high number of patients (14-18). Indeed, the analysis of data in observational studies is based on the association of clinical outcomes with mean values of measurements in several patients, which compensate for the inaccuracy of their individual results. On the other hand, the success of RCTs depends on the accuracy of the results obtained in each singular patient, whose therapy is tailored based on PFT. Misclassification of patients in terms of platelet reactivity on clopidogrel will lead to unnecessary and useless switch to more efficient drugs in unrecognized good responders and to persistency of ineffective treatment with clopidogrel in unrecognized poor responders, both contributing to the failure of PFT-guided therapy.

Our meta-analysis also shows that the results of PFT-guided anti-P2Y12 therapy do not change as a function of the number of treated patients with ACS, thus discrediting the hypothesis that the failure of the early published large-scale RCTs on PFT-guided therapy was to be ascribed to the enrolment of <50% of ACS patients (58). It is noteworthy in this respect that, in a large non-randomized intervention study of 1789 ACS patients, potentiation of antiplatelet therapy corrected HTPR, but did not improve the clinical outcomes of clopidogrel-treated patients (82).

### HTPR-Therapy

RCTs of therapies for patients displaying HTPR on clopidogrel showed that alternative antiplatelet regimens significantly decreased the risk of MACE, compared with clopidogrel. This observation contrasts with the negative results of RCTs of PFT-GT, in which the same PFTs had been used to test platelet reactivity and the same alternative therapies (prasugrel, ticagrelor, high-dose clopidogrel alone or in combination with cilostazol) had been implemented for patients with HTPR (Table 1). If the superiority of prasugrel and ticagrelor over standard dose clopidogrel is well established (19,20,83), high-dose clopidogrel (150 mg od) were considered ineffective and potentially responsible for the failure of the early, large-scale RCTs of PFT-guided anti-P2Y12 therapy (58). However, a meta-analysis of 9 RCTs and 3 observational studies demonstrated that 150 mg od clopidogrel significantly decreases MACE, compared to 75 mg od (RR=0.67, 0.48-0.94; p=0.02) (84).

The contrasting results of PFT-GT RCTs and of HTPR-Therapy RCTs further support our interpretation that the most plausible reason for the failure of PFT-guided therapy is to be found in the inadequate diagnostic accuracy of PFTs for the identification of patients with HTPR. RCTs based on HTPR-Therapy enrolled only patients with HTPR, who were randomized to alternative antiplatelet regimens or standard therapy with 75 mg od clopidogrel. The observed efficacy of prasugrel, ticagrelor or high-dose clopidogrel in reducing the risk of MACE in these RCTs actually mirrors the results of RCTs that compared the same regimens in unselected patients at risk for MACE (19,20,83,84). In contrast to RCTs of HTPR-Therapy, in RCTs of PFT-GT, only a minority of enrolled patients in the experimental arm (patients with HTPR) were treated with alternative regimens. Failure of PFTs to recognize patients with HTPR in the experimental arm would preclude them from being treated with alternative, more efficient regimens and, consequently, lead to underestimation of the clinical benefit for the overall patient population in the experimental arm. Our interpretation is also supported by the observation that, at variance with genotype-GT and PFT-GT RCTs which showed selective advantages for patients enrolled in China, the point estimates for RR of MACE were similar in HTPR-Therapy RCTs performed in China (RR=0.56, 0.43-0.74, p<0.00001) and elsewhere (RR=0.58, 0.27-1.23, p=0.16) (Table 2). Statistical significance was reached only in RCTs performed in China, due to the higher percentage of HTPR patients on standard therapy experiencing MACE (14.1% vs 3.9%) (Figure 3).

### Limitations

Some heterogeneity was detected in the included RCTs. Definition of MACE varied in different RCTs: only myocardial infarction/acute coronary syndrome and cardiovascular/total deaths were considered by all RCTs (Table 1). Few RCTs used the BARC definition for major bleeding; some RCTs used the TIMI, GUSTO or STEEPLE criteria (Table 1).

### Novelties of our systematic reviews with meta-analyses

Our results confirm those of 2 previously published meta-analyses which showed that genotype-GT improves the outcomes of patients undergoing PCI compared with unguided standard therapy, which, however included also some RCTs that based therapeutic decisions on both genotype-GT and PFT-GT and had a short follow-up (≤30 days) (50,51). In contrast, we analyzed only RCTs that were solely based on the results of genotyping and had a follow-up of at least 1 month, which we consider more adequate to assess differences in the incidence of clinical endpoints. To the best of our knowledge, our meta-analyses are the first to report the results of relative risks for clinical endpoints in RCTs of PFT-GT and HTPR-therapy, selected based on strict, pre-defined criteria.

### Conclusions

The 2022 Clinical Pharmacogenetics Implementation Consortium Guideline for CYP2C19 Genotype and Clopidogrel recommends CYP2C19 genotyping of patients undergoing PCI (85), principally based on the results of a meta-analysis (46) that, however, analyzed together genotype-GT and PFT-GT, and did not differentiate between studies performed in China and those performed elsewhere. Based on the results of our meta-analysis, we emphasize that most of the evidence of genotyping efficacy stems from RCTs in Chinese patients, in whom PFT guidance also proved effective. Evidence from our meta-analysis indicates that, in patients from countries outside East/South Asia, genotype guidance is of dubious efficacy and PFT guidance is ineffective and should therefore be discouraged.

## Data Availability

All data produced in the present study are available upon reasonable request to the authors

## DISCOLSURES

Cattaneo M., Rocchetti M. and Minardi A. have no disclosures to declare. Birocchi S. has received honoraria from Boehringer Ingelheim; Podda GM has received consulting fees from Bayer, Sanofi, Novartis, Boehringer Ingelheim; Squizzato A. has received honoraria from Daiichi Sankyo, Bayer, Pfizer, Bristol-Myers Squibb, Boehringer Ingelheim, Sanofi, Werfen, Alexion, Roche, Viatris.

